# From Spike to Seizure: Transformation or Transition?

**DOI:** 10.1101/2025.07.18.25331676

**Authors:** Thandar Aung, Aude Jegou, Patrick Chauvel

**Affiliations:** Epilepsy Center, University of Pittsburgh Medical Center, Pittsburgh, PA, USA; Epilepsy Center, Neurological Institute, Cleveland Clinic, Cleveland, OH, USA

**Author notes:** Corresponding author: Patrick Chauvel Corresponding email address Corresponding address: 9500 Euclid Ave, Cleveland, OH 44195. Co-First Author, equally contributed (Thandar Aung, Aude Jegou).

## Abstract

**Objective:** The transition from interictal discharges to ictal high-frequency activity (HFA) remains poorly understood. We investigated whether spike-associated high-frequency oscillations (Sp-HFOs) during interictal and preictal periods contribute to the emergence of ictal HFA.

**Methods:** We retrospectively analyzed the interictal to ictal transition in seizures from six patients with drug-resistant focal epilepsy who underwent stereo-EEG and subsequent surgical resection. Various interictal periods preceding seizure onset were selected for comparison. Time– frequency analysis (TFA) was used to characterize Sp-HFOs and ictal HFA. Frequency overlap was quantified using the I-Fusion metric, and linear regression assessed changes in I-Fusion values over time, with R² indicating correlation strength.

**Results:** Visual analysis of the time series revealed a preictal phase in all patients, during which brief high-frequency activity gradually emerged within spikes (Sp-HFOs), ultimately transitioning into sustained ictal HFA at the same frequency. TFA demonstrated increasing frequency similarity with time between Sp-HFOs and ictal HFA. I-Fusion values and R² coefficients rose consistently, indicating a progressive convergence in frequency content. Notably, Sp-HFOs and ictal HFA shared narrow-band frequency features within the same electrode contacts, especially in the epileptogenic zone (EZ).

**Interpretation:** Our findings support a dynamic, frequency-specific evolution from interictal Sp-HFOs to ictal HFA, suggesting that seizure onset is preceded by a gradual preparatory phase rather than an abrupt transformation. The progressive nature and spectral continuity of Sp-HFOs may reflect increasing neuronal synchrony, providing potential early biomarkers for seizure prediction and improved localization of the EZ.

## Introduction

The question of electrical markers for a focal seizure onset remains debated, despite their limited diversity. “High-frequency activity” or “fast activity” (HFA or FA), detectable by SEEG for over sixty years^1^, has shown clinical and pathophysiological relevance in numerous presurgical intracranial studies, as well as in experimental and computational modeling research^2–6^. Nevertheless, HFA in itself can hardly be taken as a distinctive, self-sufficient pattern^7,8^. Instead, HFA consistently combines with other elements such as “preictal” or ictal spikes, sharp waves, electrodecrement, and infra slow activity^6,9^. Understanding the relationships between HFA and these associated elements, particularly preictal spikes, could be a pertinent approach to elucidating how HFA is generated.

A linked question concerns the relationship between ictal onset and interictal patterns. Along with spikes or sharp waves, high-frequency oscillations (HFOs) have been recognized as part of the interictal epileptiform activities^2,3,10^. The co-localization of interictal epileptiform discharges (IEDs) HFOs with seizure onset, as marked by the emergence of HFA, has been debated, but recent evidence now strongly supports it^10^. One challenge is the difficulty in distinguishing pathological from physiological HFOs in cortical areas^11,12^, raising questions about the effectiveness of searching for epileptogenic zone (EZ) biomarkers outside the peri-ictal period.

Moreover, statistical comparisons of interictal spikes and HFOs for EZ localization have not consistently favored fast activities^13^. One reason is that not all spikes are equivalent; spiking activities have long been observed outside of the EZ^13,14^. Discriminating between “green spikes” and “red spikes” has been the focus of numerous studies^15,16^. Careful interpretation of SEEG is crucial for distinguishing primary irritative zone (IZ) from secondary IZ^17,18^ . Notably, certain electrophysiological characteristics of spike subpopulations have been shown to be specific markers of the EZ^19^. Classifying multiple interictal features has proven valuable for EZ localization, especially spikes accompanied by gamma activity, as confirmed by surgical outcomes^2^. Such findings align with research in focal cortical dysplasia (FCD), where interictal patterns often delineate the EZ more clearly than seizure onset itself^20^. Nevertheless, the precise pathophysiological relationship between interictal activities and the constituent elements of ictal onset electrical pattern has not yet been established.

The present study represents a major step forward in reconciling the disparate and global observations on the relative value of spikes and HFOs recorded under different conditions by aiming to analyze the interplay between spikes, whether interictal or preictal, and HFA during the transition from interictal to ictal discharge. Through time-frequency analysis (TFA), a focal ictal onset is characterized by narrow-band HFA that is preceded by a single or repetitive spike activity. This raises the key question: how does a low-voltage HFO arise from a high-voltage phasic discharge? Using TFA, the frequency of ictal HFA was measured and compared to the evolving frequency components of spikes (Sp-HFOs) during preictal period and at various interictal intervals before seizure onset. Our findings revealed a continuum between preictal Sp-HFO and ictal HFA, suggesting that Sp-HFOs can arise well in advance, even during interictal period, and ultimately contribute to the emergence of sustained HFA.

## Method

### 2.1 Patient selection and Criteria

With IRB approval from the University of Pittsburgh, we retrospectively reviewed six patients with focal drug-resistant epilepsy who underwent SEEG followed by resection (2020-2023). Inclusion criteria were (i) seizure onset marked by HFA preceded by “pre-ictal” spiking and (ii) a single seizure type with stereotypic semiology and electrographic features. Seizure outcome was accessed using the Engel classification^21^ at last follow-up visit. SEEG electrodes (DIXI medical) were implanted according to pre-implantation hypotheses using the Talairach method, and SEEG signals were recorded at a sampling rate of 2048 Hz (NicoletOne^TM^ EEG system).

### 2.2 Data Selection and preprocessing

To investigate the relationship between Sp-HFOs during the periods of interictal, pre-ictal, and ictal HFA, we first focused on the interictal-ictal transition period then analyzed various background interictal periods. “Ictal” was defined as the onset of sustained HFA with concomitant low-frequency suppression, and “preictal” as the period immediately preceding HFA onset. For each patient, two 60-second SEEG segments, starting 20 seconds before the onset of HFA, were selected along with four 1-minute background segments at 1 hour, 30 minutes, 5 minutes, and 2 minutes prior. SEEG electrode locations were co-registered using GARDEL^22^, and a bipolar montage was created from grey matter channels. Visual spike detection was performed independently by two reviewers (TA and PC) across all 10 segments per patient.

### 2.3 Time frequency analysis of background interictal, preictal and preictal-ictal transition

Figure 1A presents data analysis flow chart. TFA was performed on both the interictal-ictal transition and background data, focusing on high-frequency analysis using the analytic wavelet transform, as recommended by Roehri et al.,^5^ which is known for effectively handling HFOs. We applied the Derivatives of Gaussian wavelet (2-256 Hz, 8 octaves, 12 voices) for decomposition, followed by “H0 z-score” (Z_H_0__) normalization^5^, which fits a normal distribution to wavelet coefficients for standardization. For seizures, normalization parameters were computed from the 20-second window preceding the seizure. Channels showing the epileptogenic zone fingerprint pattern, characterized by distinct transient spikes followed by narrow-band HFA, accompanied by the suppression of low-frequency activity^6,23^, were selected for further quantitative analyses.

**Figure 1:**
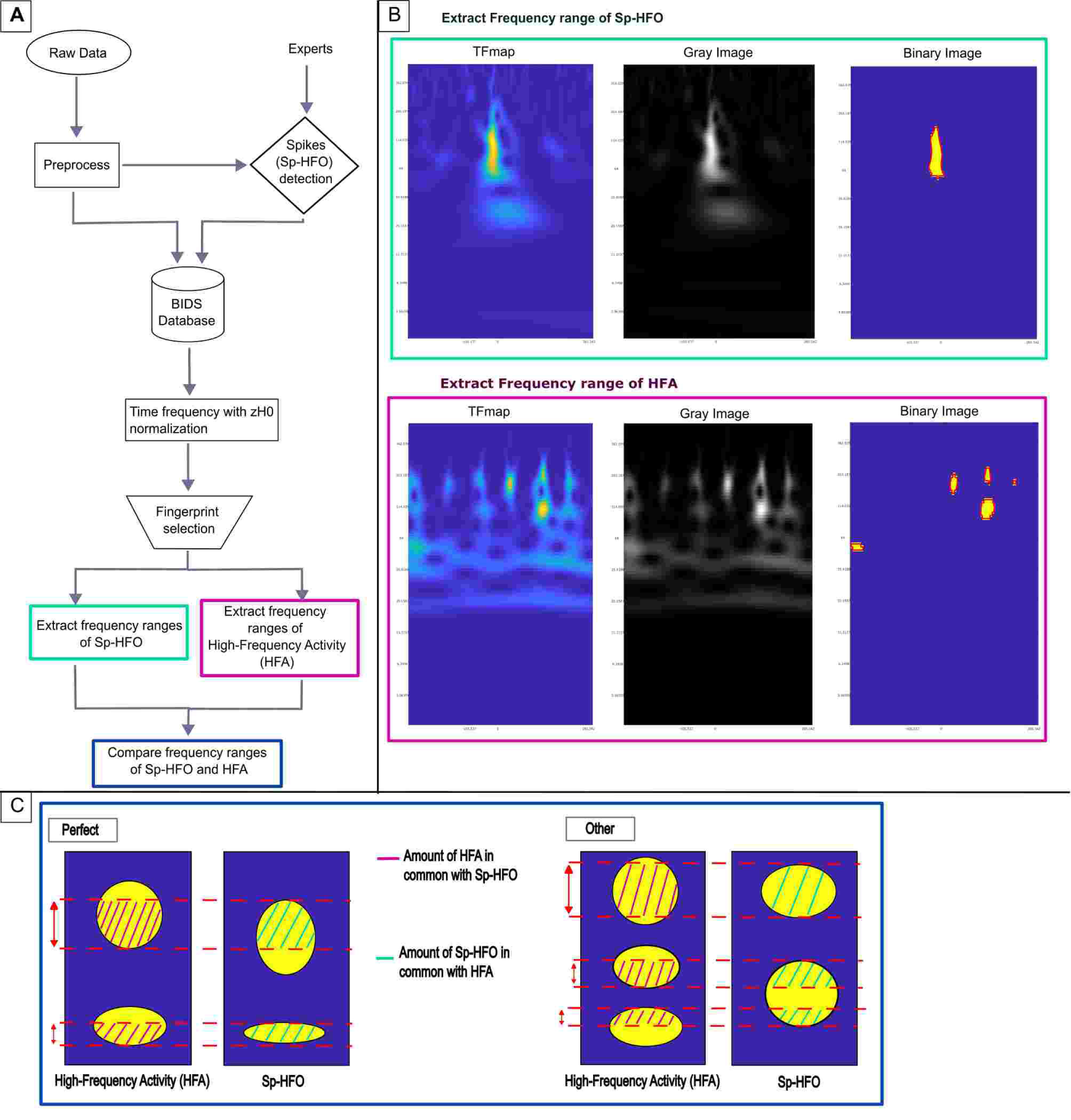
Overview of data processing workflow and frequency spectral pattern comparison method. **Panel A**: Flowchart summarizing the data analysis pipeline. Raw SEEG data segments were extracted from background intervals (60, 30, 5, and 2 minutes prior to seizure onset) and from the seizure itself. Expert reviewers (TA and PC) visually identified spikes across all segments. Data were preprocessed using bipolar montage and organized in Brain Imaging Data Structure (BIDS) format. Time–frequency analysis was computed using Z_H_ normalization. Channels exhibiting ictal “fingerprint” patterns were selected for further analysis. Frequency ranges of spike-associated HFOs (Sp-HFOs) and ictal HFA were extracted from their respective time– frequency maps (TFmaps) and compared to assess spectral similarity. **Panel B**: Illustration of the frequency extraction and comparison method. For each Sp-HFO, a 300-ms TFmap window was centered on the spike peak (green box). The grayscale image represents the TFmap, and the binary image shows thresholded high-intensity frequencies. Red contours indicate the detected frequency band edges. The same method was applied to ictal HFA windows (pink box). **Panet C**: Spectral overlap between Sp-HFOs and ictal HFA was computed and categorized into two groups: “Perfect match” (high overlap) and “Other” (partial or no overlap). The yellow blobs represent the frequency band compared to each other. The Red lines shows the edges of the common part from Sp-HFO and HFA. The pink represents the amount of HFA in common with Sp-HFO and the green represents the amount of Sp-HFO in common with HFA. (Abbreviation: Sp-HFO: High Frequency Oscillation component of the spike, HFA: High Frequency Activity)

### 2.4 Quantitative correlations of Sp-HFOs across various interictal and preictal period with HFA using I-Fusion

Selected channels exhibiting HFA at interictal to ictal transition were further analyzed for Sp-HFOs. Visually marked spikes were centered at peak amplitude and extracted in 300ms windows. Spikes were then extracted from the TF map. Corresponding TF maps were transformed to grayscale (0 to 255), binarized using a 99th percentile threshold derived from histogram-based normal distribution fitting and used to delineate dominant frequency ranges (Fig 1B).

To quantify ictal HFA, a sliding window (300ms window length with a 500ms step size) was manually set at the HFA transition position, isolating the first four windows to exclude post-HFA low frequency rebound of spiking. The same algorithm was applied to extract frequency ranges from each window of interictal and preictal period.

Frequency ranges were extracted from each channel and compared. Two parameters were computed: (1) “amount of HFA”, the proportion of overlapping frequencies between Sp-HFOs and HFA, divided by the total number of bins frequencies (bin=1Hz) in the HFA ranges, and (2) “amount of spike”, the proportion of intersecting frequencies between Sp-HFOs and HFA, divided by the total number of bins frequencies in the Sp-HFOs range (Fig 1C).

To integrate both frequency range overlap and band matching, I-Fusion (Ideal Fusion), a metric inspired by the F-measure, was developed.

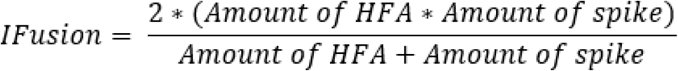

Ranging from 0 (no match) to 1 (perfect match), I-Fusion quantifies the alignment between Sp-HFOs and ictal HFA. Only Sp-HFOs with I-Fusion > 0.5 were included in further analysis to ensure meaningful match between Sp-HFOs and HFA. Sp-HFO counts were normalized per channel as a percentage of the highest count observed in each patient, with 100% indicating the channel with the highest number of Sp-HFOs.

Sp-HFO counts were normalized per channel as a percentage of the maximum count per patient. Each Sp-HFO was classified as “perfect” if it exactly matched both the frequency range and band count of ictal HFA, or “other” if only partial overlap, where Sp-HFOs and HFA shared some overlapping frequency ranges and frequency bands, was present.

### 2.5 Temporal Dynamics of I-Fusion: R-Squared Analysis

To access progressive changes in HFA from the preictal to ictal period, Sp-HFOs were chronologically ordered from the first baseline spike to the last preictal spike before HFA onset. Linear regression was applied to assess the relationship between I-Fusion values and the spike timing relative to HFA transition. The R-squared coefficient was calculated to quantify the fit of the regression model, with values close to 1 indicating a strong correlation and values near 0 suggesting a weak correlation. A positive slope indicated increasing similarity to ictal HFA as seizure onset approached.

## Results

Six patients were included in the study (Table 1): two patients with mesial temporal lobe epilepsy (patient #1 and patient #6) and four patients with neocortical epilepsy (insular-opercular (patient #2), parietal (patient # 3 and #4), and occipital epilepsy (patient #5)). Two stereotypical seizures from each patient were analyzed. In all 12 seizures, the transition between interictal and ictal activities of the seizures was investigated.

**Table 1:**
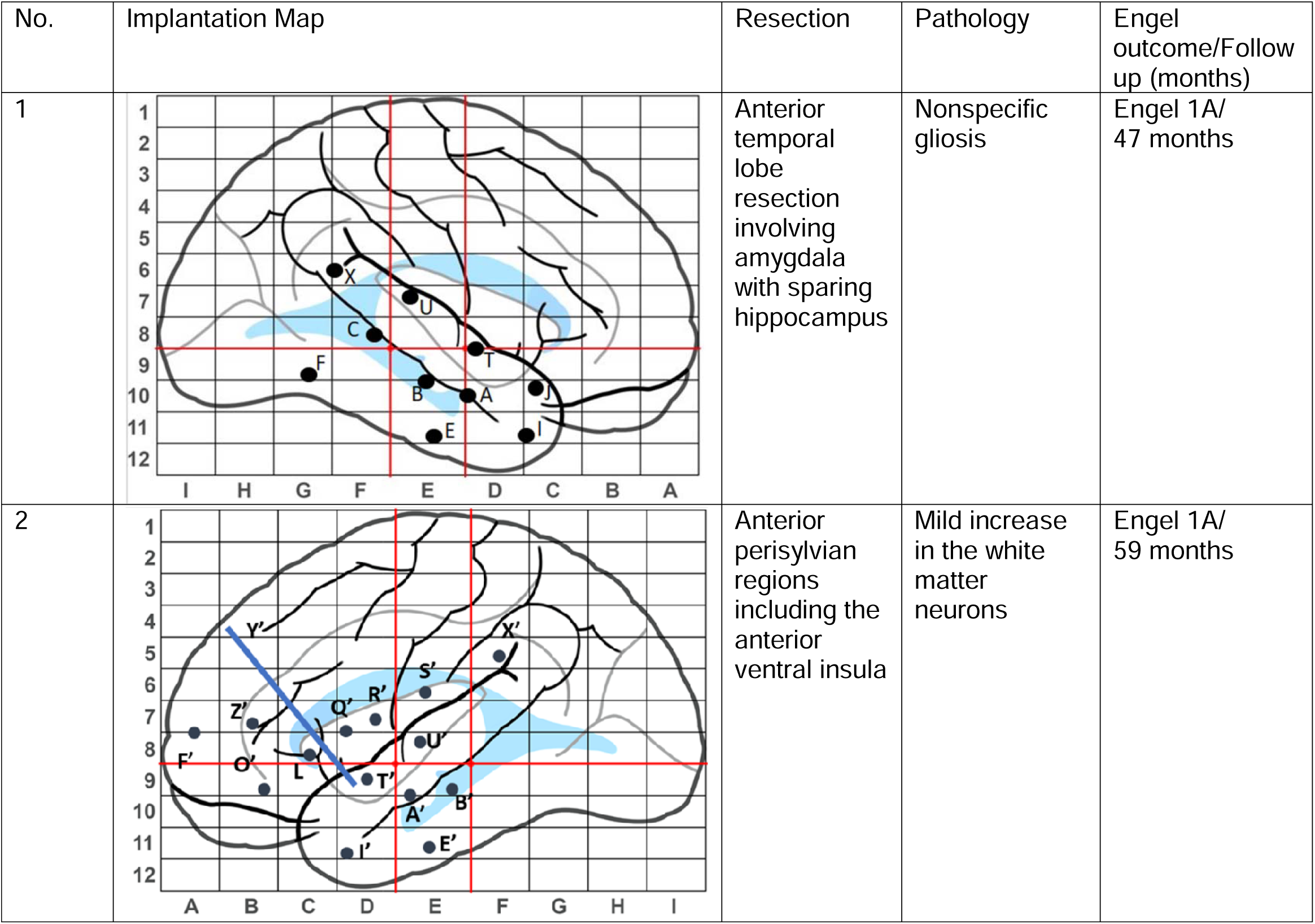
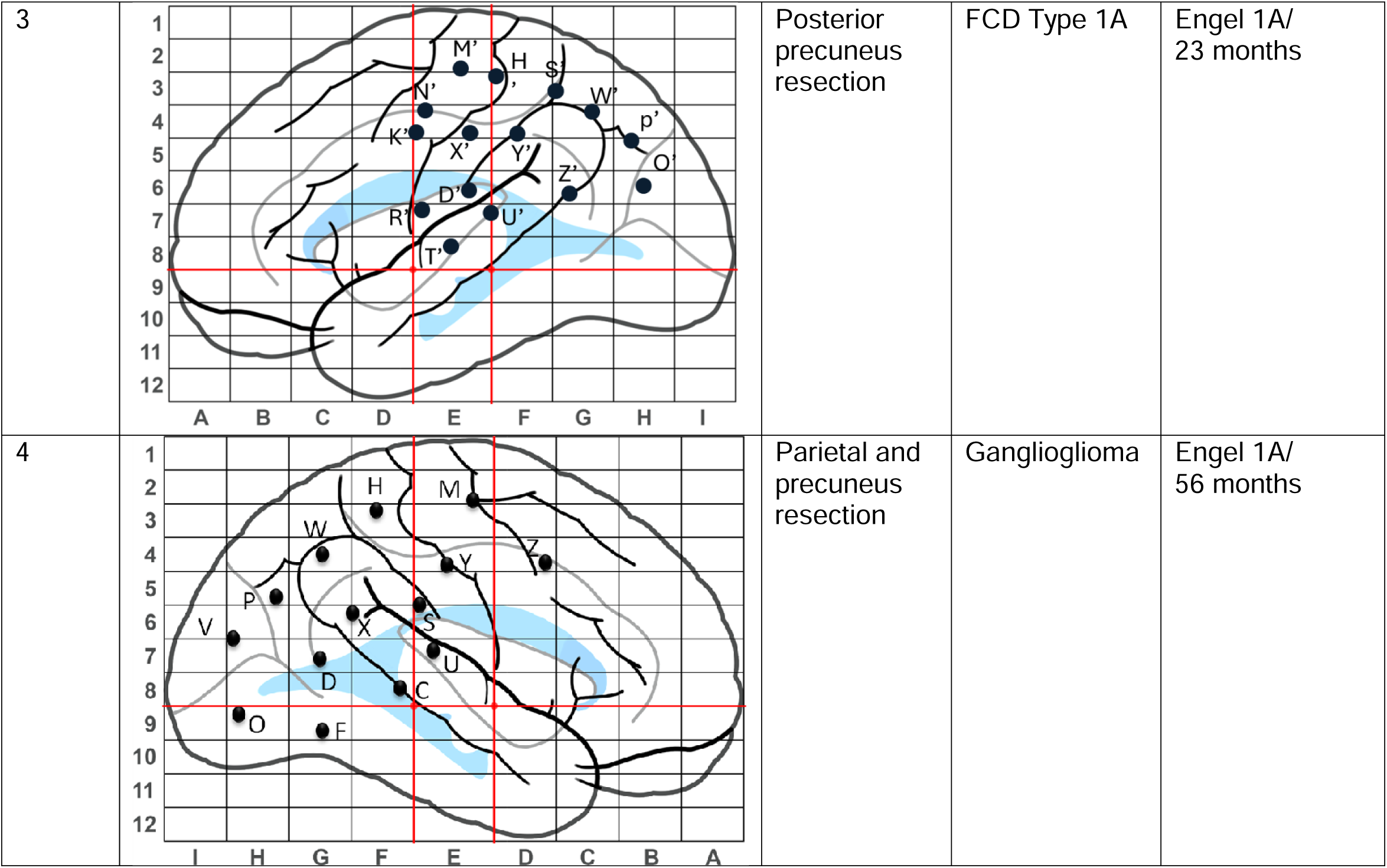
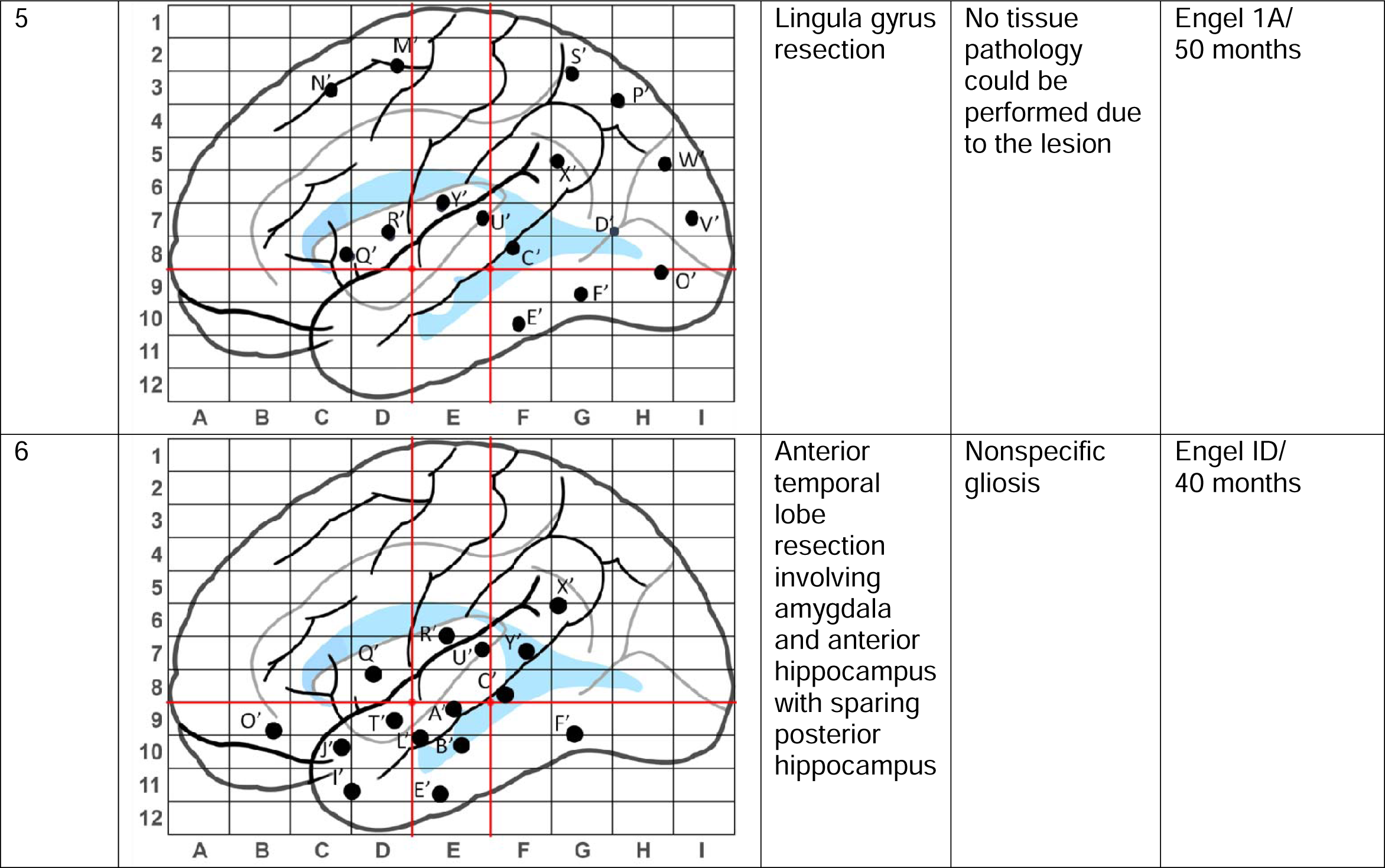
Enrolled Patient information. (Abbreviation: M: Male, F: Female)

**Patient #1**, in their 40s, had MRI-negative right mesial temporal lobe epilepsy. Figure 2A illustrates the transition from interictal to preictal to ictal states. SEEG revealed ∼8 seconds of preictal spiking, progressively synchronizing across the amygdala (A2–3), hippocampus (B2–3, C3–4), and entorhinal cortex (E1–2). Low-amplitude spikes first emerged in the amygdala and intensified with superimposed HFOs, which gradually appeared on the spike peak and descending slope. Spike amplitude and HFO prominence increased in the amygdala and hippocampus, while HFOs remained absent in EC. Sp-HFOs continued uninterrupted into ictal HFA in the amygdala and hippocampus (Fig 2B).

**Figure 2:**
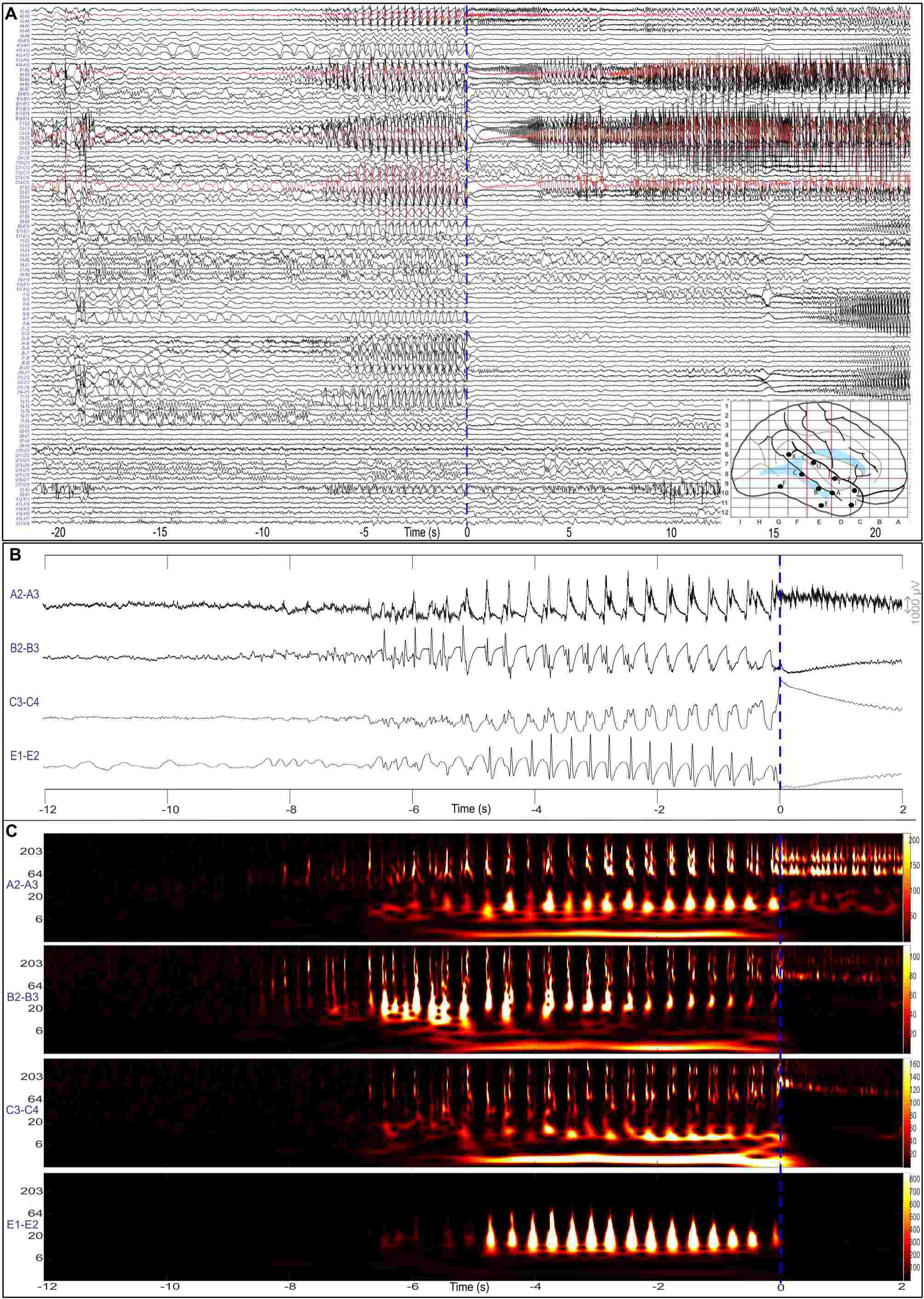
Detailed Qualitative analysis of the patient #1 stereotyped seizure using SEEG time series (A/B) and Time frequency Analysis (C). **Panel A** illustrates the qualitative assessment using visual inspection of the SEEG time series, 20 seconds before and after the ictal onset, which revealed a sequence of approximately 8 seconds of preictal spiking activity followed by 4 seconds of low-voltage fast activity. On the left-bottom corner, the schema of the SEEG montage is illustrated to explore the MRI-negative right mesial temporal lobe epilepsy. **Panel B** focuses on the time series of 4 channels of interest, taking 12 seconds before the ictal onset and 2seconds after. The spiking activity gradually became synchronous across several structures, including the amygdala (A 2-3), anterior hippocampus (B 2-3), posterior hippocampus (C 3-4), and entorhinal cortex (E 1-2). Low-amplitude spike activity was first noted in the amygdala. As the sequence progressed, the spikes became more prominent in the amygdala, accompanied by the intrusion of HFOs. At the same time, synchronous spiking was observed in the anterior hippocampus, followed by the posterior hippocampus and entorhinal cortex, all within ∼2 seconds. Sp-HFOs became prominent, appearing atop and on the descending slope of larger spikes in the amygdala. Similar findings were observed in the hippocampus but not in the entorhinal cortex. As the spiking stage progressed, both the amplitude of spikes and the prominence of HFOs increased in the amygdala and anterior and posterior hippocampus. Toward the end of this phase, spikes disappeared, whereas Sp-HFOs continued uninterrupted into the ictal sustained HFA in the amygdala. A similar observation was noted in the hippocampus, but not in entorhinal cortex. **Panel C** displays the corresponding time frequency analysis of the 14-second time series centered at the onset of HFA (12 seconds before and 2 seconds after). In the amygdala, a progressive 3-Hz spiking discharge exhibited a dual frequency pattern consisting of a low frequency component (∼ 10-20 Hz) and a higher-frequency component with two distinct bands: one centered at ∼85 Hz (range 70-100 Hz), and ∼ 176Hz (range 135-216 Hz), from –6s to 0s. The higher frequency component gradually encroached on the spike. As the spiking stage progressed, this component split into two distinct frequencies—approximately ∼85 Hz and ∼176 Hz—by around –3 seconds. The subsequent ictal HFA appeared as a prolongation of the preceding Sp-HFOs, now clearly divided into two bands. As this pattern emerged, the lower frequency band was suppressed (i.e., spike component). A similar sequence occurred synchronously in the hippocampi (anterior and posterior), though with lower frequency patterns (∼100 Hz) and less signal energy, especially in the higher-frequency components. In contrast, On the other hand, no Sp-HFOs nor ictal HFA was noted in the entorhinal cortex. Interestingly, the sustained HFA appeared to follow the last preictal spikes’ HFO component in both preictal and ictal periods were in the same frequency range (Abbreviations: HFO: High frequency oscillation, Sp-HFOs: high frequency component of spike, HFA: high frequency activity, s: second)

Figure 2C displays the TFA (14-second window centered on seizure onset), which helps disentangle the multiple frequency patterns evolving throughout the two-stage sequence. In the amygdala, a dual-band frequency pattern emerged (∼85 Hz and ∼176 Hz) against a background of 3 Hz spiking. As HFA onset approached, these frequencies intensified and separated, while lower-frequency components faded. Similar, albeit lower-amplitude, frequency patterns were observed in the hippocampus but not in the entorhinal cortex. Interestingly, the sustained HFA appeared to follow the last preictal spike’s HFO component, with both preictal and ictal periods exhibiting similar frequency range.

In A2–3, spectral continuity across the transition was evident. Figure 5A shows A1–A4 had the highest spike counts, with 25–45% I-Fusion ≥ 0.5. A2–3 had consistent perfect Sp-HFO matches, across all time intervals, and rising I-Fusion values, peaking at 0.6 during preictal period (boxplot). Figure 6A shows linear regression with R² = 0.29 in amygdala, indicating clustering of perfect Sp-HFO matches near ictal onset. This suggests that preictal Sp-HFOs increasingly resembled the frequency characteristics of ictal HFA as the seizure approached.

**Patient #2** was an adolescent with MRI-negative epilepsy localized to left anterior insular-opercular cortex. Figure 3A illustrates the evolution from interictal spikes to preictal to ictal states using SEEG time series, revealing ∼ 16 seconds of preictal spiking, progressively synchronizing across the anterior insula (L’1–2, Y’1–2), anterior cingulate (Z’1–2), and posterior orbitofrontal cortex (O’ 8-10). Initially, low-amplitude spikes emerged in AI, intensifying with superimposed HFOs, which gradually intruded on the spikes’ peak and slope. As the spiking progressed, both spike amplitude and HFO prominence increased in insula and cingulate cortex, whereas HFOs remained sparse in the PFC. Sp-HFOs then persisted uninterrupted into sustained ictal HFA from the last preictal spike in the insula and cingulate cortex, but less significantly in the orbitofrontal cortex (Fig 3B).

**Figure 3:**
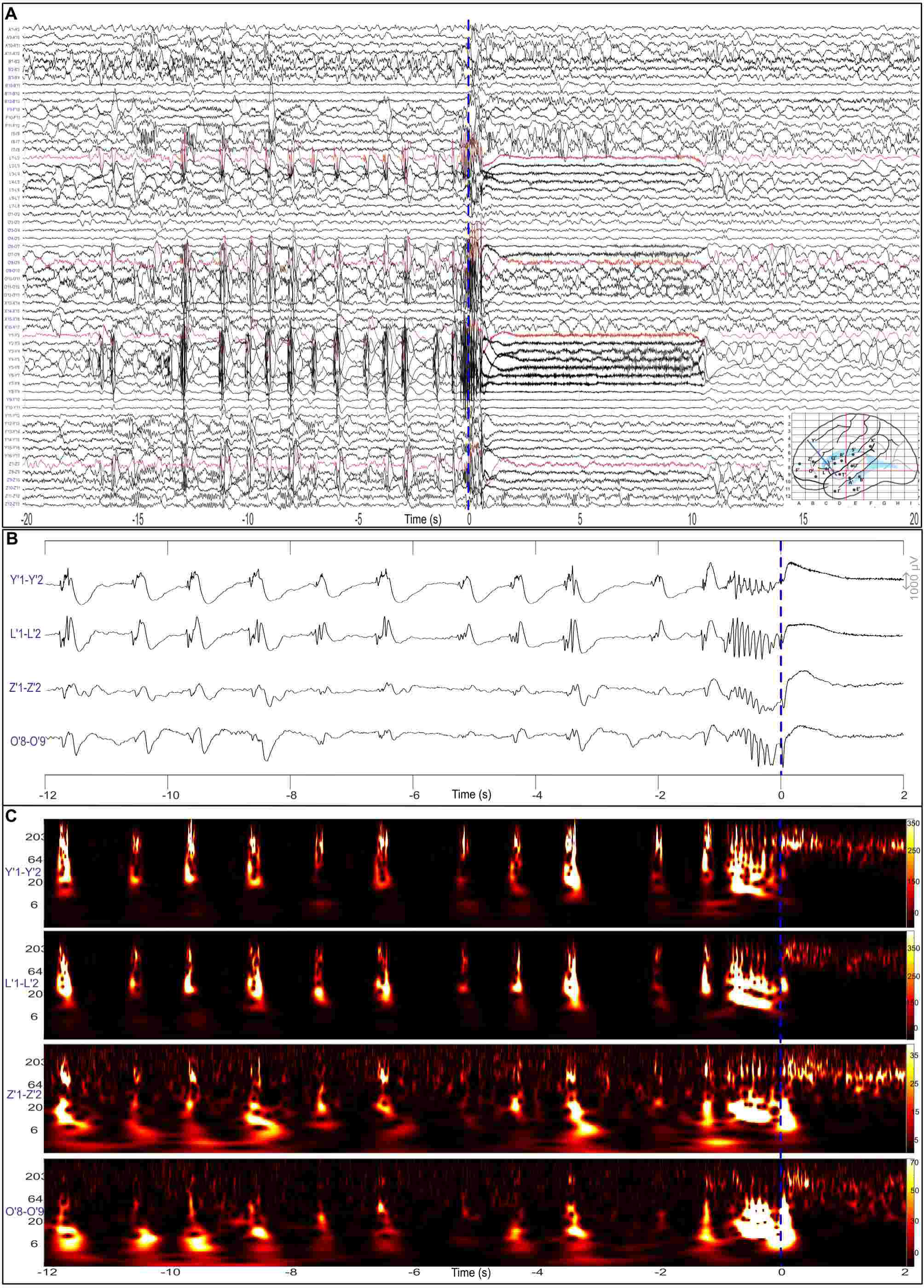
Detailed Qualitative analysis of the patient #2 stereotyped seizure using SEEG time series (A/B) and Time frequency Analysis (C). **Panel A** illustrates the transition from interictal to preictal to ictal states during the patient#2’s stereotypical seizure, 20 seconds before and after the ictal onset. On the left-bottom corner, the schema of the SEEG montage is illustrated to explore the MRI-negative left insulo-opercular epilepsy. **Panel B** focuses on the time series of 4 channels of interest, taking 12 seconds before the ictal onset and 2seconds after. Initial spikes were detected in the insular cortex (L’ 1-2 and Y’ 1-2) approximately 16 seconds before the onset of ictal HFA, synchronous with activity in the anterior cingulate (Z’ 1-2) and orbitofrontal cortex (O’ 8-9). As spiking continued, Sp-HFOs became more prominent in the insula, accompanied by increased spike amplitudes around 1 second. Sustained HFA subsequently emerged after the last preictal spike in the insula. A similar pattern was observed in the anterior cingulate cortex, while in the orbitofrontal cortex, Sp-HFOs were less prominent, and ictal HFA was absent or minimal. **Panel C** demonstrates the corresponding time frequency anaylsis, depicting a 14-second window spanning from 12 seconds before to 2 seconds after HFA onset. In the insular cortex, a progressive 1.5-Hz spiking discharge displayed a dual frequency pattern composed of low-frequency activity (∼10 Hz; range 6–12 Hz) and high-frequency components (20–203 Hz) at ∼1-second intervals. As spiking advanced, the high-frequency components intensified and separated into two bands—approximately ∼42 Hz (20–64 Hz) and ∼168 Hz (133–203 Hz) by –3.5 seconds. Eventually, the higher-frequency activity dominated, forming sustained ictal HFA, while the low-frequency component (∼10 Hz) was suppressed in the insulo-opercular cortex. A similar, though lower-intensity, sequence was observed in the anterior cingulate (Z’1–2). In contrast, the orbitofrontal cortex exhibited only rare Sp-HFOs and low-amplitude, short-duration ictal HFA. Notably, the sustained HFA appeared to follow the last preictal spike’s HFO component, with both preictal and ictal periods maintaining a consistent frequency range. (Abbreviations: HFO: High frequency oscillation, Sp-HFOs: high frequency component of spike, HFA: high frequency activity)

Figure 3C TFA shows a 14-second window centered on ictal HFA onset. In the AI, a 1.5-Hz spiking discharge with multiple frequency activities: low-frequency (∼10 Hz) and high-frequency components (∼42 Hz and ∼168 Hz), with recurring intervals of ∼1 second. As the spiking advanced, the low-frequency activity diminished, while the high-frequency components intensified and split into two distinct bands by ∼3.5 seconds before seizure onset. A similar, though weaker, spectral pattern was observed in the AC, with minimal activity in the orbitofrontal cortex. Interestingly, the sustained HFA appeared to follow the last preictal spike’s HFO component in both AI and AC, whereas ictal HFA was largely absent in the orbitofrontal cortex.

Multiple spikes across the insular-opercular and cingulate regions showed high alignment with HFA, with I-Fusion values ≥ 0.5 (Fig 5B). Y’1–2 demonstrated best balance between the number of Sp-HFOs detected and the percentage of perfect matches with ∼68% perfect matches across all interictal periods (histogram), with I-Fusion consistently above 0.65. Perfect matches occurred in >50% of background periods, with a modest, non-significant increase toward the ictal transition (boxplot). Linear regression analysis revealed a positive correlation in the Y’1– Y’2 with R² > 0.2 (Fig 6B).

**Patient #3,** in their mid-20s, had MRI-negative left parietal epilepsy. Supplementary Figure 1A shows 1-Hz spikes appeared in the precuneus and gradually organized into ∼4-Hz spiking as seizure approached. As the spiking stage progressed, spike amplitude and Sp-HFO prominence increased in precuneus (P’4–5), parieto-occipital sulcus (O’5–6), parietal operculum (S’7–8), and posterior cingulate gyrus (Y’3–4). Sp-HFOs then continued into ictal HFA in precuneus and adjacent regions (Supplementary Fig 1B).

TFA (14-second window centered on seizure onset) helped disentangle the multiple frequency patterns evolving throughout the two-stage sequence (Supplementary Fig 1C). A dual-band frequency pattern emerged (∼64 Hz and ∼114 Hz) in Channels P’4–5 and O’5–6 over a background of lower-frequency spiking around ∼20 Hz (range 11–20 Hz). As HFA onset approached, the high-frequency components (∼114 Hz) intensified, and low-frequency faded. Similar patterns were noted in parietal operculum and posterior cingulate gyrus. Sustained HFA followed the last preictal Sp-HFO, exhibiting a consistent frequency range.

Figure 4A illustrates TFA comparisons between Sp-HFOs across time periods and ictal HFA, with I-Fusion arrows indicating the degree of frequency overlap (1 = perfect match, 0 = no match) across frequency range of 2–256 Hz. Channels P’4–5 and P’5–6 exhibited the highest percentage of Sp-HFOs matching the ictal frequency range, with I-Fusion values exceeding 0.5 in more than 60% of detected spikes (Fig 4B). The average I-Fusion for perfect Sp-HFOs across all background intervals was ∼0.7, increasing to ∼0.8 during the preictal period, indicating stronger spectral overlap near ictal onset. Channel P’ 4-5 showed elevated I-Fusion values in the preictal period (∼0.7) compared to the 5-minute background (∼0.55) (boxplot). Linear regression analysis revealed five channels—P’2–3, P’3–4, P’5–6, P’6–7, and P’7–8—with positive correlation between I-Fusion values and time, each with an R² ≥ 0.2 with a maximum reached at 0.45, indicating a strong correlation with the ictal frequency evolution (Fig 6C).

**Figure 4.**
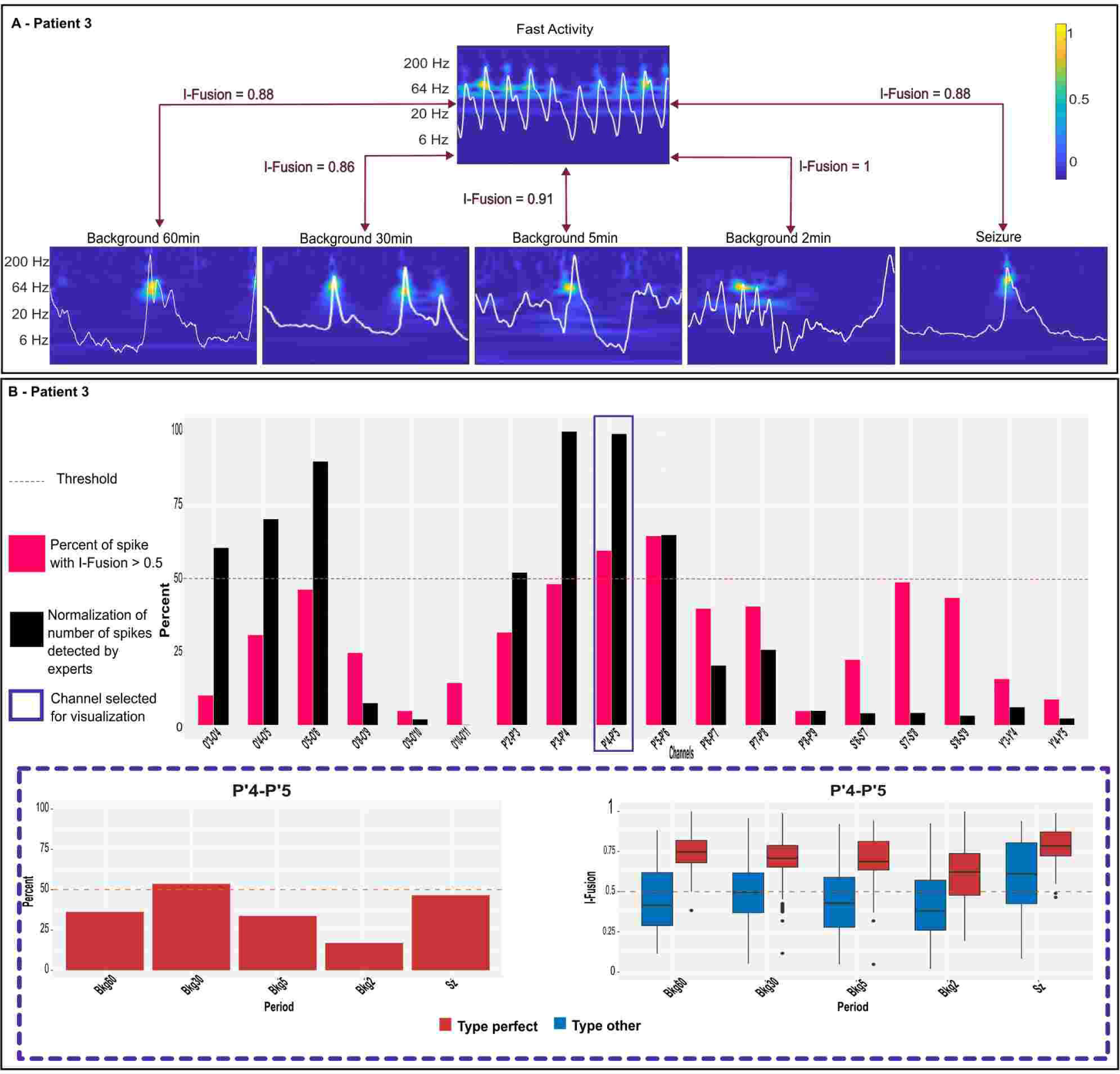
Spectral overlap between Sp-HFOs and ictal HFA in Patient #3. **Panel A:** Time– frequency maps (TFmaps) illustrating Sp-HFO events from five preictal time points (60, 30, 5, and 2 minutes before ictal onset, and preictal) compared to ictal HFA (top row). Each TFmap represents a 300-ms window (2–256 Hz) with SEEG time series overlaid in white; color scale reflects signal magnitude (blue: low; yellow: high). Red arrows denote I-Fusion values, a metric of spectral similarity (0 = no match, 1 = perfect match). **Panel B**: Histogram displays, by channel, the percentage of Sp-HFOs with I-Fusion ≥ 0.5 (pink), indicating high spectral overlap with ictal HFA. Black bars show normalized Sp-HFO detection rates across channels. The purple-highlighted channel exhibits consistent high I-Fusion values across time points. The inset histogram shows temporal distribution of perfect matches of selected channel P’4-5; the adjacent boxplot illustrates I-Fusion distribution by different time periods and Sp-HFO match types in the same channel. Highest I-Fusion averages occurred during the preictal period (∼0.8), suggesting increased spectral convergence prior to seizure onset. (Abbreviation: Bkg60: Sixty minutes before the seizure onset, Bkg30: Thirty minutes before the seizure onset, Bkg5: Five minutes before the seizure onset, Bkg2: Two minutes before the seizure onset and SZ: at the onset of the ictal high frequency activity)

**Patient #4**, in their 40s, had a 28-year history of parietal epilepsy post-oligodendroglioma resection. Supplementary Figure 2B shows initially disorganized, continuous spiking with slowing activity in the precuneus (W1–2), which later became more organized and intensified with superimposed HFOs. As the spiking stage progressed, synchronized spike activity was noted across mid-cingulate gyrus (Z 5-6), post-cingulate gyrus (S3-4) and parietal cortex (P 7-8), accompanied by increasing spike amplitude and greater HFOs prominence in all regions. Toward the end of this phase, Sp-HFOs continued uninterrupted into the ictal sustained HFA.

TFA revealed a broadband high-frequency pattern (36–114 Hz) in Channel W1–2 (Supplementary Fig 2C). As HFA onset approached, the high-frequency components intensified and persisted into the ictal period as sustained HFA following the last preictal spike’s HFO component, while the lower-frequency spike component gradually disappeared. Similar, lower-amplitude patterns were observed in the parietal and cingulate cortex.

Most parietal and cingulate channels exhibited high I-Fusion values exceeding 75%, with W1–2 maintaining ∼50% perfect matches and a slight increase during the preictal period (Fig 5C, histogram). The boxplot showed the highest proportion of Sp-HFOs aligned with ictal HFA in W1–2, with a mean I-Fusion value of ∼0.8, consistent across interictal periods. Linear regression identified a lower R² (∼0.1), indicating stable Sp-HFO patterns with minimal correlation to the ictal period (Fig 6D).

**Figure 5:**
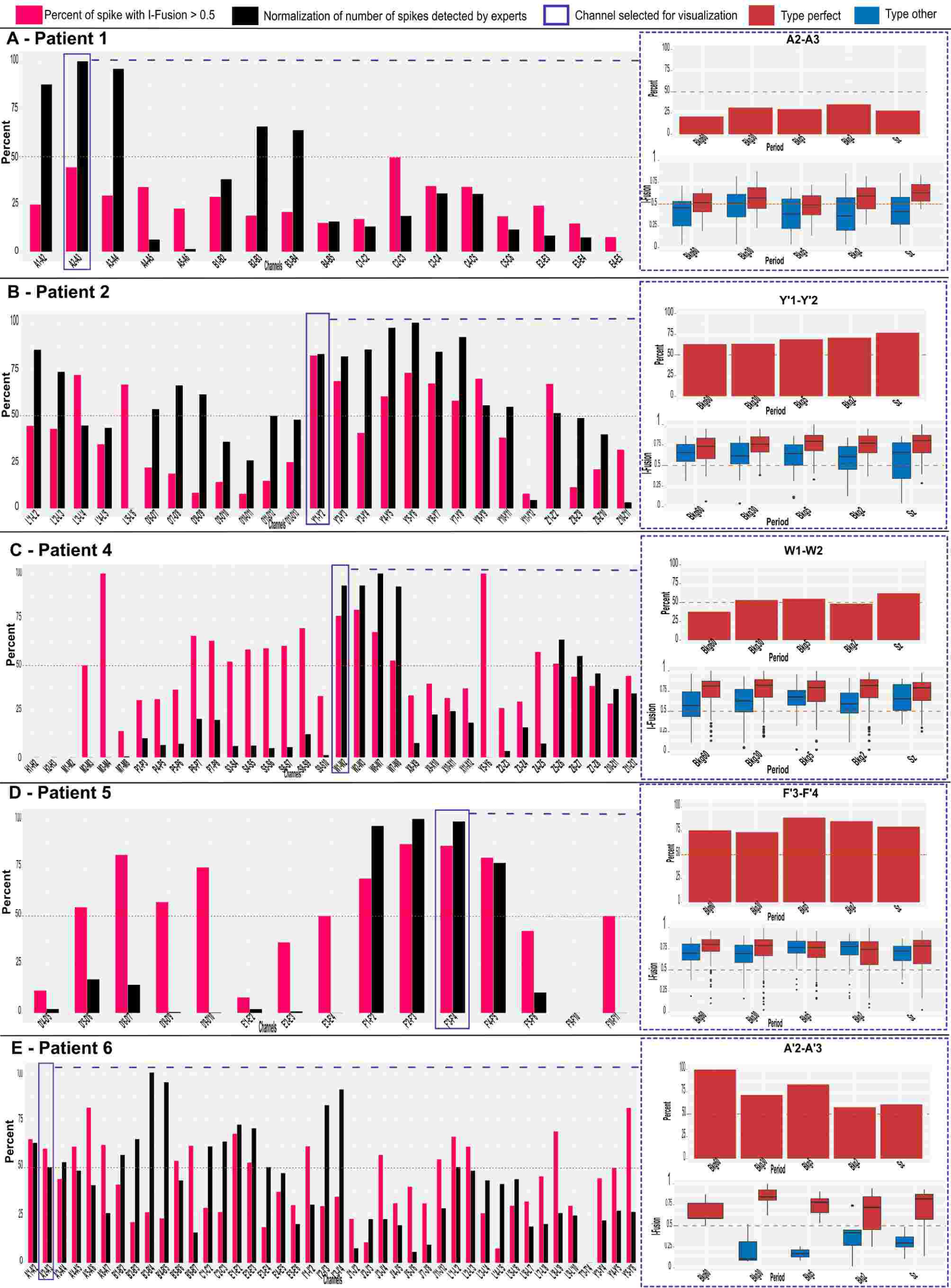
Channel-wise distribution and temporal evolution of spectral similarity between Sp-HFOs and ictal HFA across patients. Each panel represents data from a single patient. Left-side histograms display, by channel, the percentage of Sp-HFOs with I-Fusion ≥ 0.5 (pink), indicating spectral similarity to ictal HFA. Black bars represent normalized Sp-HFO detection rates across channels (100% = most Sp-HFOs detected; 50% = half that number). A 50% threshold (orange line) aids in identifying high-yield channels. The right-side insets highlight representative channels (purple boxes), showing the temporal distribution of “perfect” matches by period (left histogram) and corresponding I-Fusion distribution for perfect vs. non-perfect matches (boxplot). **Panel A: Patient 1:** All channels exhibited <50% of Sp-HFOs with I-Fusion ≥ 0.5. Channel A2– A3 (amygdala), with the highest detection rate, showed ∼45% spectral matches. Perfect matches remained <50% across all periods, with I-Fusion values slightly increasing toward seizure onset (max ∼0.6 in the preictal period). **Panel B: Patient 2:** Multiple channels in the insular-opercular region (Y’, L’) demonstrated >60% spectral matches. Channel Y’1–2 showed the optimal balance of match rate and event count, with perfect matches >50% across all periods and increasing over time. I-Fusion values averaged ∼0.75 for perfect matches and >0.5 for others. **Panel C: Patient 4:** Most channels showed >50% spectral overlap. In precuneus (W1–2), perfect matches approached 50% across all periods with a slight increase preictally. I-Fusion values remained high (mean ∼0.8 for perfect, ∼0.7 for others) and stable across time. **Panel D: Patient 5:** Most channels exhibit >= 50% of matching. However, only mesial F channels (lingula gyrus) have many Sp-HFO detected. F’3–4 showed ≥75% perfect matches consistently. I-Fusion distributions were uniform across time, averaging ∼0.75 for both perfect and non-perfect types. **Panel E: Patient 6:** Most channels showed ∼50% spectral matches. Channel A’2–3 (amygdala) exhibited ≥75% perfect matches in all periods, with 100% in the 60-minute background (likely due to low event count). Boxplots revealed high I-Fusion for perfect matches (>0.75), while non-perfect matches remained <0.5. (Abbreviation: Bkg60: Sixty minutes before the seizure onset, Bkg30: Thirty minutes before the seizure onset, Bkg5: Five minutes before the seizure onset, Bkg2: Two minutes before the seizure onset and SZ: at the onset of the ictal high frequency activity, Sp-HFOs: high frequency component of spike, HFA: high frequency activity)

**Figure 6.**
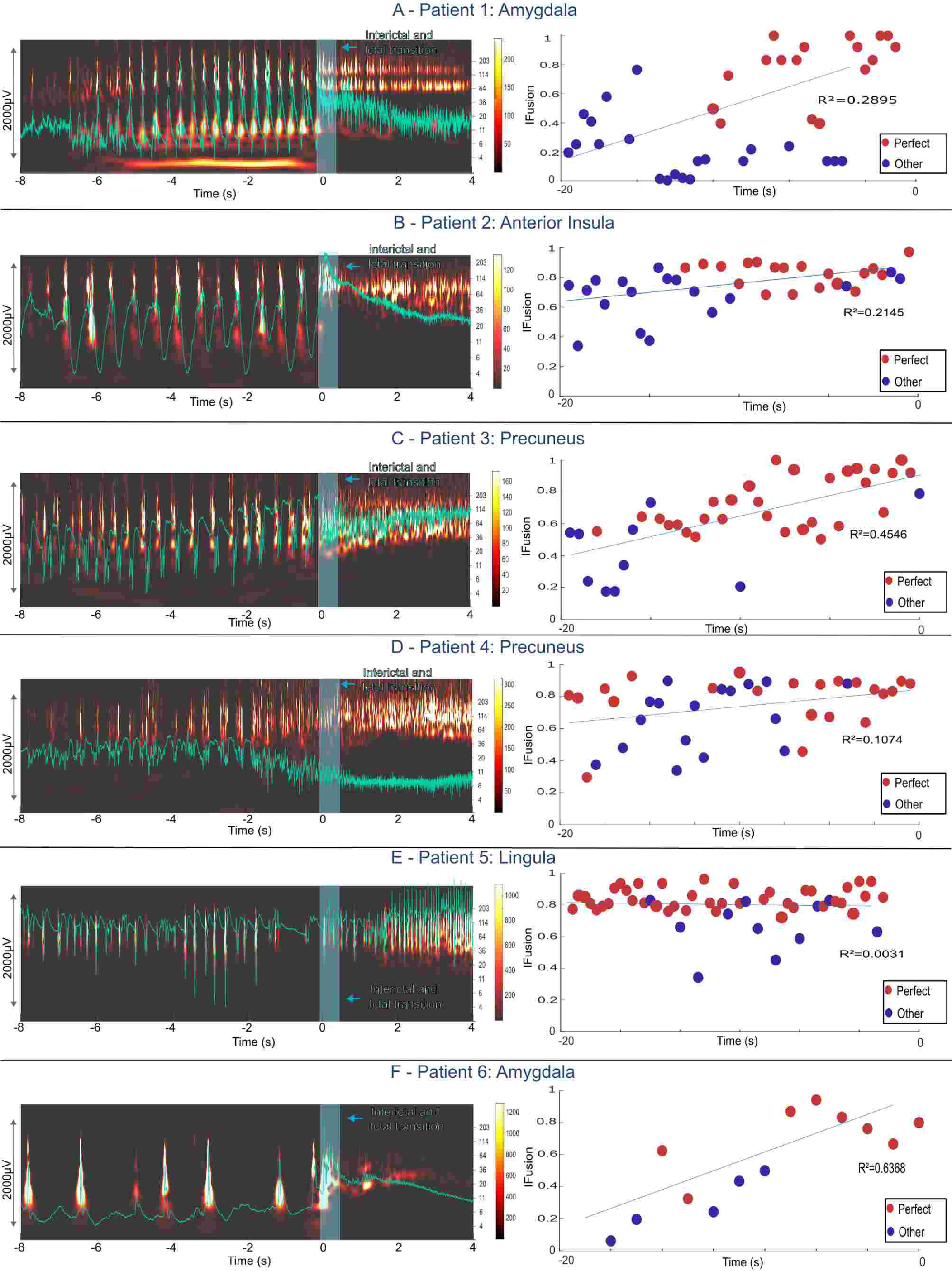
Time–frequency analysis and correlation of Sp-HFO spectral similarity to ictal HFA across patients. Each panel corresponds to one patient. Left panels show time–frequency maps (2–256 Hz, logarithmic scale) over a 12-second window centered on seizure onset (time 0) with SEEG time series overlaid in green. Signal magnitude is indicated by color (black = low; red = moderate; white = high). Light blue boxes highlight the transition from preictal to ictal states. Right panel displays scatter plot of R² values calculated from linear regression of I-Fusion scores across time to quantify the correlation between Sp-HFO spectral features and ictal HFA onset. The red dot represents the « perfect » matches while the blue dot represents the « other » matches. **Patient 1**: Amygdala exhibited a modest positive correlation (R² = 0.29), indicating a gradual increase in spectral alignment as seizure onset approached. **Patient 2**: Anterior Insula showed a consistent temporal relationship with ictal HFA, with a moderate R² > 0.2. **Patient 3**: Precuneus exhibited the highest R² reached 0.45, indicating strong preictal-to-ictal spectral convergence. **Patient 4:** Precuneus demonstrated a low R² (∼0.1), suggesting stable but non-progressive Sp-HFO patterns. **Patient 5:** Lingula did not reveal a significant temporal trend; R² values remained low, consistent with a stable Sp-HFO-HFA relationship across time. **Patient 6:** Amygdala exhibited the highest R² (> 0.6), reflecting a strong correlation between Sp-HFO spectral features and ictal HFA.

**Patient #5**, in their mid-30s, was a male with a 22-year history of focal MRI-negative occipital lobe epilepsy. Supplementary Figure 3B shows left lingual cortex (F’1–4) had continuous interictal spikes increasing to 7–8 Hz, with superimposed HFOs. HFOs became prominent and lengthened before ictal HFA. As the seizure progressed, Sp-HFOs became more prominent and prolonged in F’1-4. Approximately 2 seconds before the ictal high-frequency activity (HFA) transition, the HFOs lengthened, and the spike frequency increased, leading to the persistence of the HFO component into the ictal HFA transition.

TFA complemented these findings by revealing a broadband HFO (30–203 Hz) in F’3–4 (Supplementary Fig 3C). As the seizure approached, the higher-frequency band (36–203 Hz) persisted and intensified, transitioning into ictal HFA within the same frequency range, while the lower-frequency band (10–20 Hz) disappeared. Notably, no preictal Sp-HFOs were detected in the EC (E’1–2), occipital cortex (O’12–13), or POS (D’5–6), and no clear ictal HFA was observed during the transition.

In the I-fusion analysis, the F’ electrode exhibited that 75% of spikes were classified as perfect matches across all periods, with an average I-fusion value of 0.8. This indicates a strong correlation between Sp-HFOs of the interictal and preictal, with HFA (Fig 5D). Notably, the linear regression did not reveal significant changes in I-fusion values over time during the preictal period (Fig 6E).

**Patient #6**, in their mid-20s, had a 24-year history of MRI-negative left medio-basal temporal lobe epilepsy. Supplementary Figure 4A illustrates synchronous preictal spikes occurring at a rate of 1 Hz, at around ten seconds before the ictal HFA. As the seizure progressed, Sp-HFOs became more prominent in amygdala (A’2-3), with synchronous spikes and Sp-HFOs noted in the hippocampus (B’ 3-4, C’ 2-3), but not in EC (E’1-2). As spike activity subsided, Sp-HFOs continued without interruption into low gamma FA within the amygdala and hippocampus, but not in the EC. (Supplementary Fig 4B)

TFA revealed persistent low gamma components (∼20 Hz, 15–25 Hz, high intensity; ∼36 Hz, 30–40 Hz, lower intensity) along with an intense low-frequency band (8–25 Hz) that disappeared at ictal onset. The ∼20 Hz and ∼36 Hz bands persisted and transitioned into ictal HFA in the amygdala, with similar patterns in the hippocampus and EC. While Patient #6 demonstrated a shorter transition period compared to patient #1, the stereotyped evolution from Sp-HFOs to ictal HFA remained consistent—even when the dominant frequencies fall within the lower gamma range, as is often seen in mesial temporal lobe epilepsy. (Supplementary Fig 4C)

In the I-fusion analysis, most channels exhibited ∼50% Sp-HFO alignment with ictal HFA, with A’2–A’3 standing out by showing ≥75% perfect matches across all periods and reaching 100% during the 60-minute background. One of the explanations for the 100% may be due to the low number of detected Sp-HFOs (Fig 5E). The boxplot revealed high I-Fusion values for perfect matches (>0.75), while non-perfect types remained below 0.5. Figure 6F illustrates (A’2–A’3) showing a strong positive correlation (R² > 0.6), with perfect matches clustering closer to the ictal period.

In conclusion, our study, using I-Fusion values and R² correlations, provides insights into the relationship between preictal Sp-HFOs and ictal HFA during the interictal-to-ictal transition. We observed a progressive correlation of preictal Sp-HFOs over time, reflected by increasing R² values approaching the ictal onset. Moreover, our analysis suggests that interictal HFO components of spikes or IEDs show preparatory activity related to ictal HFA, as evidenced by increasing I-Fusion rates and perfect matches.

## Discussion

To figure out how the ictal high frequency activity arises at the onset of focal seizures is the central question in this study. Since the new trend of presurgical investigation with SEEG, multiple studies have concluded that the “low-voltage fast activity” (LVFA) at seizure onset is the most robust marker for epileptogenic zone^8,24,25^. However, this statement came from statistical analysis studies taking surgical outcome as ground truth^26–29^. Such reference makes sense only when surgical resections are of limited size. After identification of HFOs as interictal epileptiform discharges, the relationship between high frequency interictal activity features and epileptogenic zone was also investigated based on statistical co-localization^10,30^. Then, a debate ensued on which of the spikes or of the HFOs were the best marker for epileptogenic zone from the interictal period^31^. There is now no question that both contribute to seizure triggering. Given that LVFA per se is not a reliable marker of an epileptogenic zone unless it is made of narrow high frequency band and combined with preictal spiking and low frequency suppression in the same site, the conditions of its emergence had to be studied in the context of this complex interictal to ictal transition.

A consistent time sequence was identified across all patients in this study. In a preictal phase (immediately before HFA occurrence), there was a change in the interictal activity structure. Brief bursts of high frequency activity gradually appeared and encroached upon the spikes (Sp-HFOs) before leading to sustained HFA at the ictal transition. The HFO component of preictal spikes and the ictal HFA in the epileptogenic zone channels fell within the same frequency range, indicating that the HFA generation is likely to be a continuation from the preictal phase rather than a sudden distinct and disruptive phenomenon. Among studies that questioned the relative specificity of interictal spikes vs HFOs in predicting the EZ location, some of them identified a peculiar pattern made of the combination of spikes and HFOs: “spike-gamma rates in wakefulness were associated with a high probability of achieving seizure freedom.”^10,19^

Our study confirms the interest of this association and reveals the dynamic features of a “preparatory” phenomenon. Given the prolonged spectral features of the HFA following the pre-ictal Sp-HFOs, the build-up mechanism of this phenomenon needed to be explored. The fact that the spikes and the HFOs, as well as the Sp-HFOs, could be detected and identified with SEEG macroelectrodes, denotes a high degree of synchrony among their generating neuronal assemblies. In particular, the narrow-band aspect of the HFOs is a signature of such synchrony. As the spike-nested gamma oscillations observed in the current and previous studies support an interdependence of spikes and HFOs, and the ictal HFA escapes from the preictal Sp-HFOs at the same frequency, is the ultimate synchronization subserving HFA develops only during the preictal spiking or is it already present during the interictal period, and is there an evolution of the phenomenon with time?

Therefore, how does this interplay between the spikes and the HFOs give rise to this HFA escaping from the preictal spiking stage at the same frequency? How does the sp-HFO structure evolve with time? To answer these questions, we undertook a comparative analysis of the frequency contents of interictal spikes and HFA at ictal onset by using Ideal fusion method (I-Fusion), then R²-squared analysis. I-Fusion is a dynamic measure of the frequency alignment between Sp-HFOs and HFA, offering insights into how closely the interictal activity matches the ictal frequency range. In addition, time evolution of the I-Fusion was essential to estimate with the R²-squared analysis.

Evidence of an alignment gave us a dynamic view of how closely the preictal activity matches the frequency content of the impending seizure, leading to identify the traits of a preparatory phase. The R²-squared analysis, on the other hand, measured the strength of the correlation between the preictal Sp-HFO and ictal HFA across time, revealing how these two activity patterns evolve together building up to the seizure.

The I-fusion method was effective in capturing a preparatory phase of seizure onset. Across the six patients, the I-fusion values demonstrated varying degrees of alignment between Sp-HFOs and HFA at seizure onset, highlighting distinct preparatory shifts in neural dynamics. Interestingly, in some epileptogenic zones like in amygdala and temporal pole, high I-fusion values remained consistent, indicating a robust relationship between Sp-HFOs and ictal HFA. As the ictal period approached, the I-Fusion values generally increased, suggesting a gradual shift toward preferential ictal frequencies. This trend was consistent in most of the patients, with some of them showing more distinct and pronounced preictal shifts. The lack of a clear preparatory shift in patient 5 resulted from a continuous activity of interictal spikes at the F’1-2 and F’3-4 locations, whose spectral characteristics were already the same as at seizure onset. This explained why the R²-squared analysis did not document a consistent preparatory trend throughout the preictal period. Each of the interictal Sp-HFOs was spectrally similar to seizure onset. This was not surprising in a FCD type II case. This demonstrates that localization of EZ in similar cases can be based upon interictal activity spectral content. This is an interictal fingerprint.

The difference in evolution of “perfect vs other” with time between patient 5 and the other patients (Fig 6) illustrates the existence of a gradual tidying up of the relationship between the spike and the fast activity which could hitherto present some fluctuation. Figure 6 also shows that there is a plateauing trend of “perfect” alignment at the end, meaning that the last Sp-HFOs are also more rigidly aligned to each other. Quite interestingly, the spectral pattern of the Sp-HFOs is similar to the SWR (Sharp Wave Ripple) supporting memory consolidation in the hippocampus^32^. In this model, the generation of the gamma frequency band is explained by a mechanism of concatenation of several SWRs^33^. The time-frequency plots in Figure 6 effectively show such a concatenation of high-frequency activities at the interictal to ictal transition between Sp-HFOs and HFA.

This study sheds new light on the genesis of the epileptogenic zone fingerprint (EZF)^6^. Defined by the combination of preictal spiking, narrow band HFA and suppression of low frequency activity in one brain “zone”, EZF can be considered as a valid marker of an epileptogenic zone, and a guidance for epilepsy surgery as a consequence. However, repetitive spiking. HFA, or suppression can also be recorded in isolation at seizure onset and the question of their relevance is debated ^24,34^. All the previous studies were undertaken with SEEG where a sampling bias is inherent to the method. The diverse seizure onset patterns described in those studies, which are made of various associations of isolated or repetitive spikes with low voltage activities of multiple frequency ranges, could well be due to sub-optimal electrode placement with respect to the epileptogenic zone. The current study shows how the EZF originates from a gradual development of a high frequency oscillatory activity initially nested within certain interictal spikes, before its eventual liberation at seizure onset.

As hypothesized earlier^6,23,35^, fast spiking, parvalbumin-positive inhibitory interneurons are likely to be involved in the generation of the chirping HFA at seizure onset. After a seminal experimental study demonstrating preservation of perisomatic inhibition of pyramidal neurons in pilocarpin and kainate epilepsy models, a neural mass model approach supported a failure of slow dendritic inhibition and preservation (or exacerbation) of fast perisomatic inhibition as the mechanism responsible for the emergence of high frequency activity at the onset of focal seizures^36^ . This hypothesis was ultimately confirmed through microelectrode recordings in experimental models^37–40^ . In particular, a causal relationship between the high-frequency discharge of PV+ interneurons and the HFA was clearly evidenced^16,41^. Then, neural mass modeling showed that the combination of feed-forward and feedback connections in a specific range of strength connectivity between pyramidal cells and PV+ interneurons was the best condition for generation of HFA and explained its chirping aspect^42^ .

Could a similar mechanism account for the sp-HFO and its gradual transformation into HFA? Among the abundant literature about HFOs, few studies addressed possible common mechanisms for interictal spike and nested HFO. In slices or isolated brain preparations, microelectrode recordings of entorhinal cortex showed that interictal spikes have distinct intracellular correlates from the preictal spikes: while the preictal spikes correlate with inhibitory interneurons discharges, the interictal spikes correlate with bursts of action potentials superimposed on a paroxysmal depolarization shift in pyramidal cells ^43,44^. So, the role of inhibitory interneurons would be prominent only at the preictal-ictal transition. However, to the best of our knowledge there is no phenomenon like interictal spike-HFO in experimental epilepsy. Therefore, the best model for it so far could be the hippocampal pathological ripples (pripples)^45^. Even though the relative contribution of pyramidal cells and inhibitory interneurons is unclear, several studies show that their discharges are overlapping in time and the HFO component of the local field potential corresponds to high-frequency bursts of action potentials in pyramidal cells^32^ . However, distinctive features of SWRs and interictal epileptiform discharges (IEDs) were clearly demonstrated: in IED, the initial event was fast firing of PV+ basket cells leading to a depolarization block leading to increased firing of pyramidal cells maximal at the peak of the IED and the peak of the HFO power^46^. Many recent papers identified a gamma-spike sequence^19,47^ that could reflect the same mechanism.

What is the significance of the gradual alignment of high frequency in the evolution of interictal discharge and its transition to ictal activity? The spectral aspect of the Sp-HFOs as a narrow band indicates a high level of synchrony in a circumscribed network. The Molaee-Ardekani model supports the hypothesis that network frequency tuning depends on the relative strength of excitatory/inhibitory connections in the pyramidal/fast inhibitory neurons couple^42^ (Fig 5 and Fig 6). Variation of this functional connectivity with time until adjusting to the best epileptogenic system frequency is a likely explanation. During the interictal Sp-HFOs stage, the two intermingled but distinct graphoelements might result from an alternate discharge of pyramidals and inhibitory interneurons. During the HFA stage, where the phasic slow sp has disappeared and the resonant frequency has been reached, the underlying mechanism could be a pure synchronous discharge of the fast inhibitory interneurons silencing the pyramidals^40,48^ followed by their depolarization block, or a coupled discharge of both preceding their decoupling and the post-inhibitory rebound.

## Data Availability

All data produced in the present study are available upon reasonable request to the authors

## Acknowledgement statement (including conflict of interest and funding sources)

We sincerely thank our colleagues in the Epilepsy Division at the Department of Neurology, UPMC, including physicians and EEG technologists, for their invaluable collaboration, which made this work possible. We are especially grateful to Dr. Jorge Gonzalez-Martinez for his skilled implantation of depth electrodes for SEEG—a critical contribution to this study. Additionally, Dr. Gonzalez-Martinez performed surgery on all patients included in this research.

## Conflict of Interest Disclosure

The authors declare no conflicts of interest and received no external funding for this study.

## Data Sharing

The data that support the findings of this study are available on request from the corresponding author. The data are not publicly available due to privacy or ethical restrictions.

## Abbreviations

EZ: epileptogenic zone
EZF: epileptogenic zone fingerprint
FA: fast activity
FCD: focal cortical dysplasia
HFA: high-frequency activity
HFO: high frequency oscillation
Sp-HFOs: spike-associated high-frequency oscillations
IZ: irritative zone
LVFA: low-voltage fast activity
SEEG: Stereoelectroencephalography

## Author Contributions

Dr. Patrick Chauvel served as the corresponding author and led the study design, data interpretation, and manuscript preparation. Dr. Thandar Aung and Aude Jegou contributed equally to data collection, statistical analysis, and critical revision of the manuscript. All three authors were involved in drafting and revising the manuscript and approved the final version

## Notes

### Competing Interest Statement

The authors have declared no competing interest.

### Funding Statement

This study did not receive any funding

### Author Declarations

IRB of the University of Pittsburgh approved this retrospective study.

