## Supplementary figures and images for "From Spike to Seizure: Transformation or Transition?"

### Supplementary Figure 1

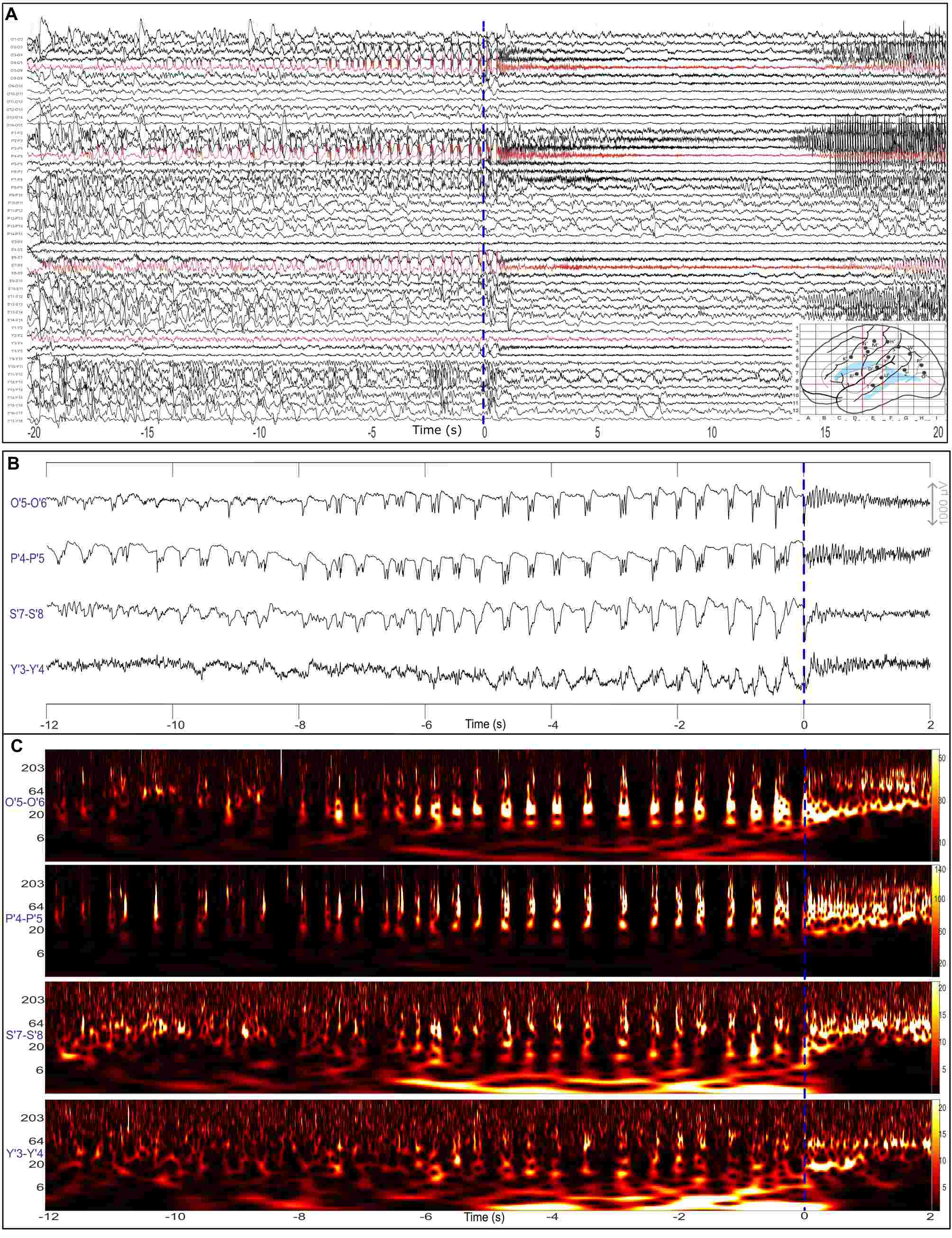

### Supplementary Figure 2

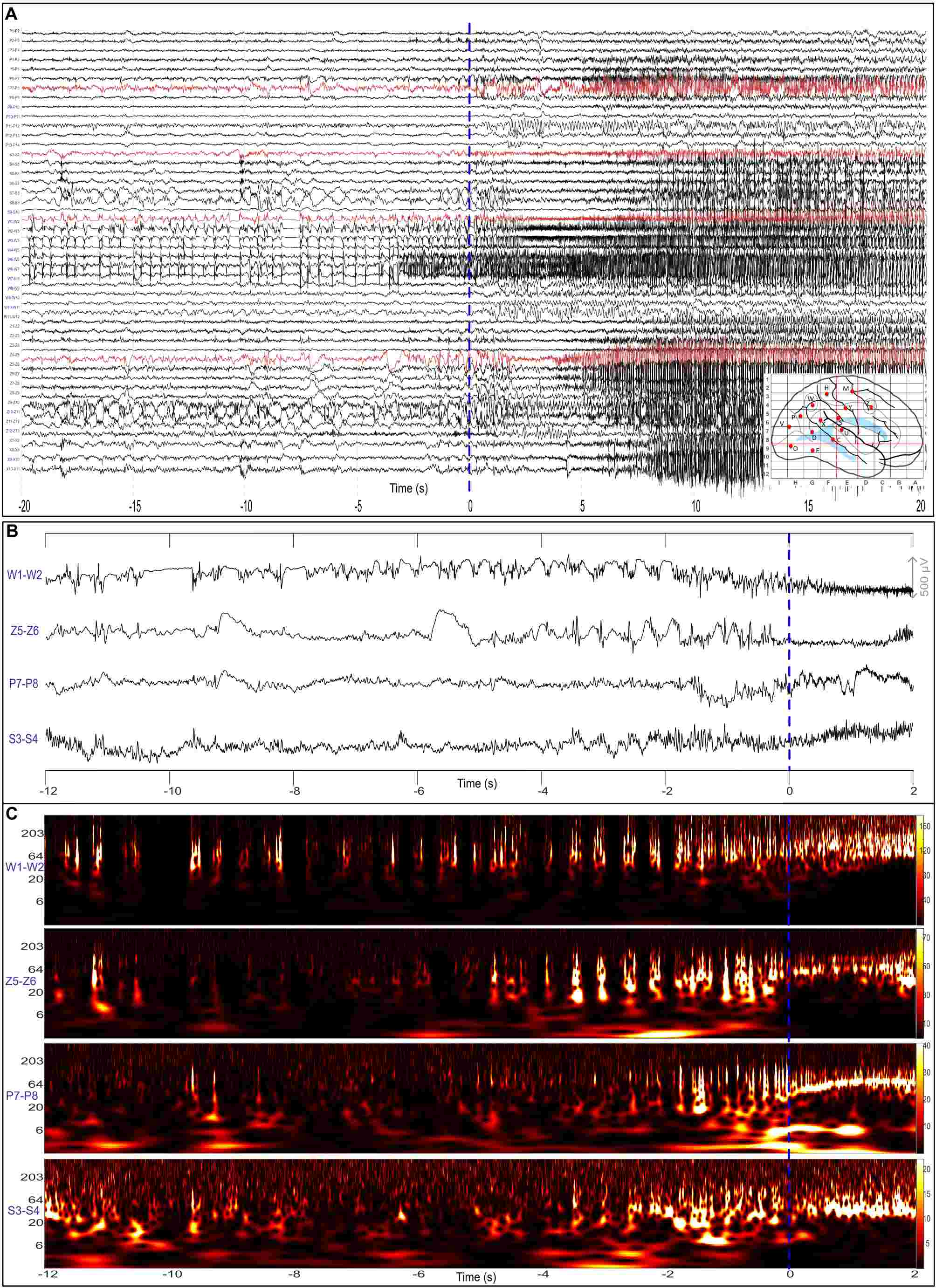

### Supplementary Figure 3

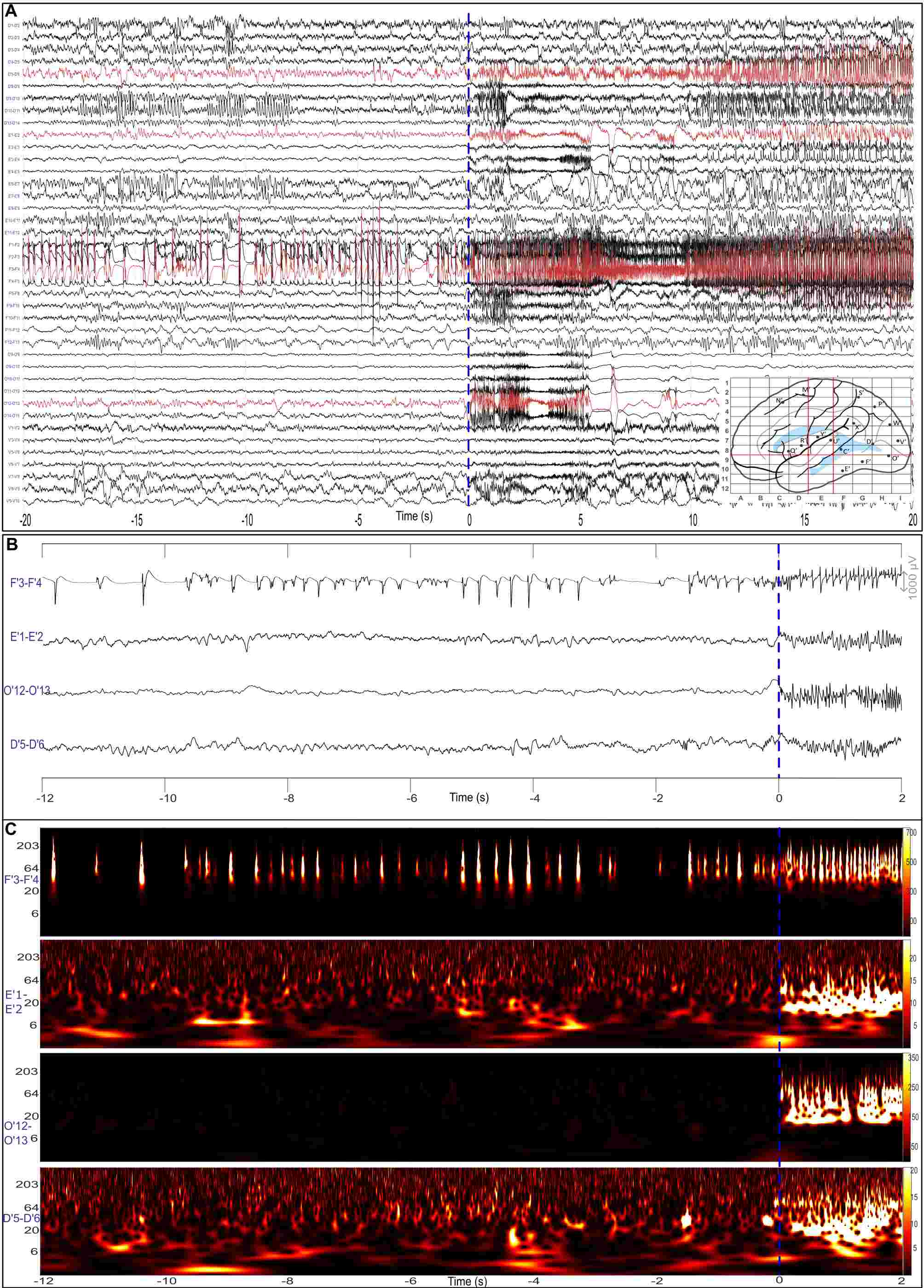

### Supplementary Figure 4

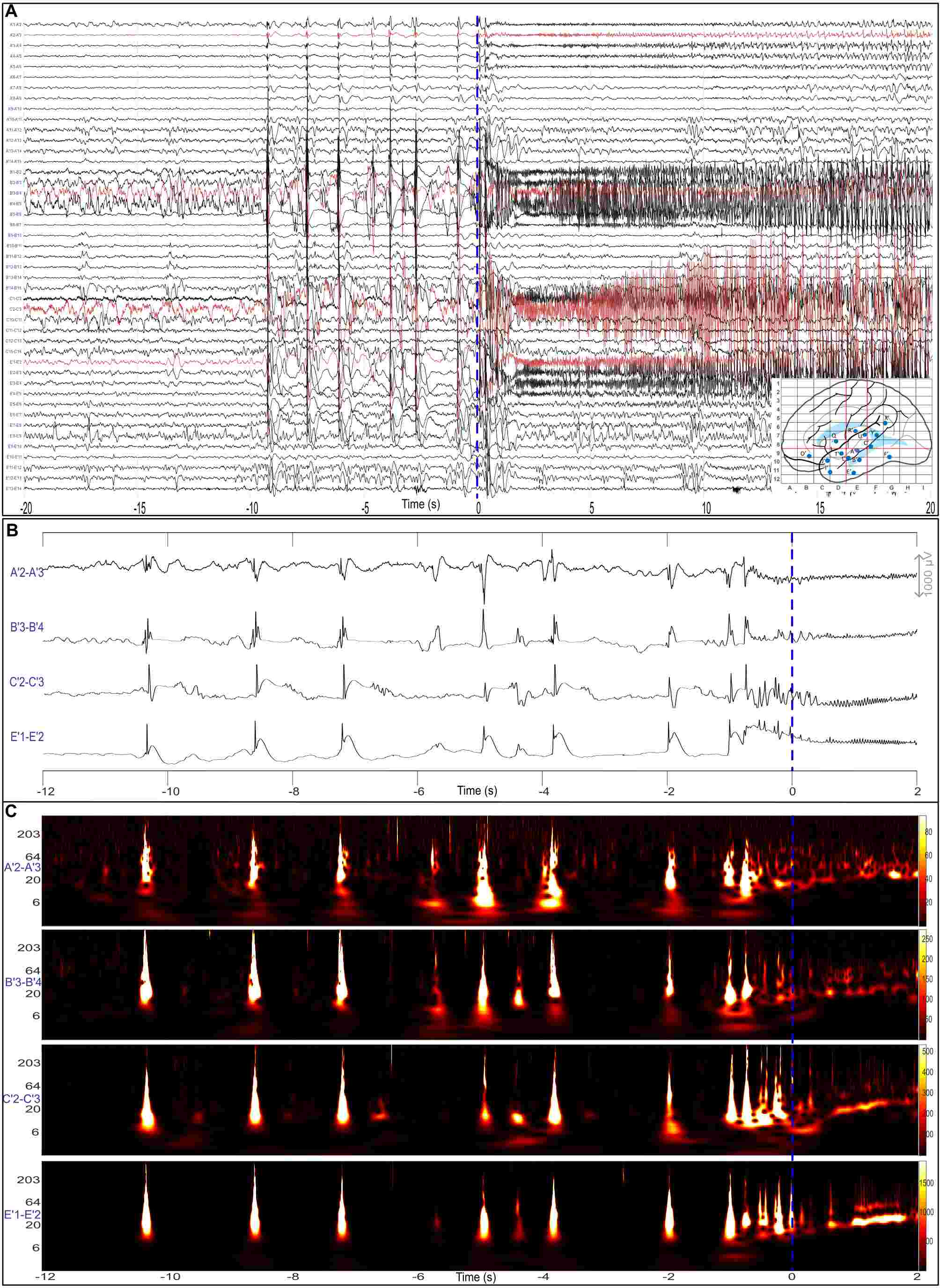
